# Evaluating social and spatial inequalities of large scale rapid lateral flow SARS-CoV-2 antigen testing in COVID-19 management: An observational study of Liverpool, UK (November 2020 to January 2021)

**DOI:** 10.1101/2021.02.10.21251256

**Authors:** Mark A. Green, Marta García-Fiñana, Ben Barr, Girvan Burnside, Christopher P. Cheyne, David Hughes, Matthew Ashton, Sally Sheard, Andrew Geddes, John Rankin, Iain E. Buchan

**Affiliations:** Senior Lecturer in Health Geography, Department of Geography & Planning, University of Liverpool, Liverpool, UK; Professor of Health Data Science, Department of Health Data Science, University of Liverpool, Liverpool, UK; Professor in Applied Public Health Research, Department of Public Health and Policy, University of Liverpool, Liverpool, UK; Senior Lecturer in Biostatistics, Department of Health Data Science, University of Liverpool, Liverpool, UK; Research Associate, Department of Health Data Science, University of Liverpool, Liverpool, UK; Lecturer in Health Data Science, Department of Health Data Science, University of Liverpool, Liverpool, UK; Director of Public Health, Liverpool City Council, Liverpool, UK; Professor of Modern History, Department of Public Health and Policy, University of Liverpool, Liverpool, UK; Chair in Public Health and Clinical Informatics, Department of Public Health and Policy, University of Liverpool, Liverpool, UK

## Abstract

**Objective:** To explore social and spatial inequalities in uptake and case-detection of rapid lateral flow SARS-CoV-2 antigen tests (LFTs) offered to people without symptoms of COVID-19.

**Design:** Observational study.

**Setting:** Liverpool, UK.

**Participants:** 496 784 residents.

**Intervention:** Free LFTs to all people living and working in Liverpool (6^th^ November 2020 to 31^st^ January 2021).

**Main outcome measures:** Residents who received a LFT, residents who had multiple LFTs, and positive test results.

**Results:** 214 525 residents (43%) received a LFT identifying 5557 individuals as positive cases of COVID-19 (1.3%) between 6^th^ November 2020 and 31^st^ January 2021. 89 047 residents had more than one test (18%). Uptake was highest in November when there was military assistance. High uptake was observed again in the week preceding Christmas and was sustained into a national lockdown. Overall uptake and repeat testing were lower among males (e.g. 40% uptake over the whole period), Black Asian and other Minority Ethnic groups (e.g. 27% uptake for ‘Mixed’ ethnicity) and in the most deprived areas (e.g. 32% uptake in most deprived areas). These population groups were also more likely to have received positive tests for COVID-19. Spatial regression models demonstrated that uptake and repeat testing were lower in areas of higher deprivation, areas located further from test sites and areas containing populations less confident in the using Internet technologies. Positive tests were spatially clustered in deprived areas.

**Conclusions:** Large-scale voluntary asymptomatic community testing saw social, ethnic, and spatial inequalities in an ‘inverse care’ pattern, but with an added digital exclusion factor. COVID-19 testing and support to isolate need to be more accessible to the vulnerable communities most impacted by the pandemic, including non-digital means of access.

**What is already known on this topic:**

- Testing asymptomatic individuals with rapid lateral flow SARS-CoV-2 antigen devices detects the most infectious individuals who otherwise would have been unaware they were likely to infect others.
- Liverpool (UK) conducted the world’s first whole population, open-access, voluntary asymptomatic testing programme for COVID-19 management.
- The impacts of such testing on inequalities are unknown.

**What this study adds:**

- Testing uptake was lower, and test positivity was higher, among deprived populations, Black Asian and other Minority Ethnic groups and areas classified as having low Internet use.
- Population-wide asymptomatic testing programmes need to account for social, spatial, and digital access issues in their design, communication and delivery to minimise inequalities in outcomes.

## Introduction

The spread of Severe Acute Respiratory Syndrome Coronavirus-2 (SARS-CoV-2), resulting in coronavirus disease 2019 (COVID-19), has been unprecedented in rapid global spread and impact on society. The difficulty in containing COVID-19 is in part due to asymptomatic cases making it difficult to monitor and prevent [1]. We define asymptomatic cases here as individuals who are infected and either display no symptoms during their infection, have symptoms that are mild and not necessarily identifiable as COVID-19 (i.e., pauci-symptomatic or subclinical), or are pre-symptomatic (i.e., individuals with replicating SARS-CoV-2 in the period before they develop symptoms, when they may be infectious). One recent study estimated that at least 50% of COVID-19 cases may have been contracted from asymptomatic individuals [2], although the prevalence of wholly asymptomatic individuals has been estimated at 20% [3].

In response to the pandemic, the UK Government and Liverpool public health authorities piloted free rapid lateral flow SARS-CoV-2 antigen testing (LFT) for people living or working in the City of Liverpool, UK [4]. Liverpool had the highest prevalence of COVID-19 at the time of planning in early November 2020. The objective was to catch cases early and break potential chains of transmission. The pilot was deployed rapidly with the assistance of the British Army and was widely advertised across different media. The pilot was extended by request of Liverpool’s public health teams, moving from a ‘mass’ (i.e., trying to test whole populations) to a SMART (Systematic Meaningful Asymptomatic Repeated Testing) approach, through targeted testing and outreach to neighbourhoods, occupations or groups at high risk (e.g., care homes) [5]. The pilot was intended to generate policy evidence on the performance, uptake and impacts of rapid asymptomatic testing. Evaluating population-level interventions is key to understanding what works, why and for whom, both for tackling the current pandemic and improving societal resilience to future infections. Equity of testing is also important as COVID-19 recovery plans look to rapid asymptomatic testing as a means of re-opening social and economic activities while containing the transmission of SARS-CoV-2, in concert with vaccination.

The impacts of large-scale COVID-19 testing on social and spatial inequalities are unknown. All COVID-19 testing in the UK is optional and the initial month of the Liverpool pilot encouraged all adults to “let’s get tested”. Downstream interventions that rely on individual agency for engagement often exacerbate existing inequalities [6,7]. For instance, uptake of self-testing technologies for HIV is lower for Black African ethnic groups [8]. Poor health literacy, mistrust of government, lack of free time to access services, concerns about insecure income or the inability to work from home and therefore self-isolate in the event of a positive test may all disproportionately influence disadvantaged or vulnerable groups to not get a test. With deep inequalities in COVID-19 outcomes evident in the UK and globally by level of deprivation, ethnic group, and geography [9,10], testing strategies and support of people to isolate are likely to further impact on these inequalities.

This study explores the social and spatial inequalities in the uptake and outcomes of large-scale rapid testing in Liverpool for people without symptoms of COVID-19.

## Methods

### Data

Person-level pseudonymised records were accessed using the Combined Intelligence for Population Health Action (CIPHA; www.cipha.nhs.uk) data resource. CIPHA was established in March 2020 to improve population health management for the 2.6m population of Cheshire and Merseyside (UK). It includes person-level linked anonymised records across NHS, local government, social care, administrative and public health information systems.

Our study is divided into four distinct periods reflecting the evolution of the pilot: (i) Initial ‘mass testing’ pilot period with military support (6^th^ November to 2^nd^ December 2020); (ii) Christmas period (3^rd^ December to 4^th^ January 2021), when Liverpool was one of two regions placed in Tier 2, with fewer restrictions on movement and economic activities than the rest of the country; (iii) return to national lockdown (5^th^ January to 31^st^ January 2021); (iv) the whole period (6^th^ November 2020 to 31^st^ January 2021). We selected all records that were identified as LFT for these periods. Three outcome variables were defined. First, the number of people who had LFT was used to provide an indicator of uptake (i.e., the proportion of the population that had received at least one test during the time period, selecting only their first test by time period). Second, we calculated the number of people who received multiple tests (i.e., selecting their second test by time period) to identify the proportion of people who had multiple tests. Finally, we calculated the proportion of all tests by time period that were positive.

CIPHA records included age, sex and ethnic group which were used to identify demographic patterns in uptake. Missing data were low other than for ethnic group (Appendix A). Following data linkage and selecting ethnicity from repeated tests, 9.8% of individuals had missing ethnicity records. Ethnic group was imputed by polytomous regression using an individual’s age and the ethnicity profile of their neighbourhood of residence. Addresses of individuals were matched to Lower Super Output Areas (LSOAs) to provide geographical location. LSOAs are small neighbourhood zones (∼1500 people). Records were aggregated to LSOAs (n=298) to allow for analysis of geographical patterns.

To provide context for geographical patterns, we matched LSOAs to their most recently available external data on key population, social and spatial determinants of testing uptake. Official mid-year (2019) population estimates by age were used to provide denominators for uptake and account for age profiles of areas [11]. Index of Multiple Deprivation (IMD) 2019 was used to measure level of neighbourhood deprivation to identify social inequalities in uptake patterns [12]. We used deprivation score for analytical models, and present summary statistics by Liverpool quintiles (to measure city-based inequalities) and national quintiles (to allow for wider comparisons as Liverpool is a highly deprived city). The proportion of university students in an area, using data from the 2011 Census, was included to account for targeted testing across Liverpool’s universities. Whether a LSOA contained a care home or not was included, using data from the Care Quality Commission (CQC), to account for targeted testing in care homes. The Internet User Classification (IUC) 2018 was selected as a proxy for confidence in using the Internet and related digital inequalities [13]. The multidimensional measure classifies areas based on their access to Internet-related infrastructure, frequency of use, and online behaviours, with descriptions of area types in Appendix C. This was due to the reliance on Internet enabled technologies for advertising the pilot, registering for tests (walk-in tests were also accepted) and receiving test results.

We only consider this variable for the uptake outcome variables and not positivity, as we did not hypothesize that digital inequality would consistently affect likelihood of a positive test. Sensitivity analyses also considered an alternative measure of Internet use (see Appendix C). Finally, we estimated the street network walking distance (km) for each postcode to the nearest test site and calculated the average distance for each LSOA to account for accessibility issues that may have affected uptake. This distance was calculated at the mid time point of each of the three periods of the pilot, as the test sites that were available varied across the study period. We did not consider this variable for analysing positivity, as we did not hypothesise it would influence likelihood of a positive test.

### Statistical analyses

Summary statistics and visualisations are used to investigate how uptake, multiple testing rate and positivity varied by age, ethnicity, area deprivation, geography over time.

We use a spatial regression framework to explore how our outcomes varied with the area-based factors outlined above, whilst adjusting for age, sex and ethnicity of test recipients. To account for spatial autocorrelation we used a Besag, York, and Mollié (BYM) model [14]. This Bayesian Hierarchical Poisson model accounts for the spatial nature of our data that would otherwise violate assumptions in standard regression frameworks. A separate model is fit for each outcome (modelling persons for number of tests and multiple tests, and tests for positivity) and stratified by time period (resulting in 12 models). For each spatial model, we used an indirect standardisation approach to adjust for the age, sex and ethnic profile of the test recipients. First, we estimate the expected count for each outcome in each LSOA, by applying the Liverpool-wide age, sex and ethnic group specific rates for each outcome to the population estimates for each age, sex and ethnic group within each LSOA. We then included the log of these expected counts as an offset in the regression model, with the observed number of people who had a test, people who had multiple tests or number of positive tests in each LSOA as the outcome. Our area-based measures outlined above were independent variables to estimate how the relative probability of each outcome varied across these measures adjusting for age, sex and ethnicity. We also plot the predicted relative rate (observed/expected) estimated for each LSOA from our models. Models were fit using Integrated Nested Laplace Approximations (INLA) [15].

Since we only have data on people who were tested, we focus on small area patterns in testing outcomes. Due to the ecological nature of our analyses and limited ability to make inferences about individuals, we also undertook two sensitivity analyses (Appendix D) using the data on individual records for people who got a test within a binomial multi-level regression framework (individuals nested within LSOAs). First, we investigated the likelihood of an individual having had more than one test. Second, we examine the likelihood of each individual having had a positive test.

All analyses were conducted using R (version 3.6.2). All analytical code is available at https://github.com/markagreen/asymptomatic_testing_evaluation.

### Patient and Public Involvement

Local community groups have been involved in the design and delivery of LFTs in Liverpool. There was no direct patient and public involvement in this analysis.

## Results

Since the introduction of asymptomatic testing in Liverpool, 43% (n = 214 525) of residents took 425 793 LFTs identifying 5557 likely infections or positive tests (1.3%) (Table 1; see Appendix B for descriptive statistics stratified by time period). 18% (n = 89 047; 42% of all individuals who got tested) of Liverpool’s population had multiple tests over the study period. More females (46%) than males (40%) accessed testing over the study period. Working age adults were more likely to have been tested (including 50% of residents aged 35-64), although the age group ‘15-34’ were over-represented by university students due to targeted testing during the pilot (Appendix Figure B1). There was lower test uptake among Black Asian and other Minority Ethnic (BAME) groups, especially among ‘Mixed’ (27%) and ‘Other’ (28%) ethnic groups. The percentage of positive tests was higher among ‘Black’ (2%) and ‘Other’ (3%) ethnic groups. Inequalities were observed by neighbourhood deprivation, with residents of the most deprived areas having both lower uptake (32% for most deprived vs 53% least deprived Liverpool quintiles) and a higher percentage of tests that were positive (1.8% for most deprived vs 1.1% least deprived Liverpool quintiles). This social gradient was present across all the time periods (Appendix B), but flatter in the national lockdown.

**Table 1:** **Summary statistics of the three outcome measures for the whole period of analysis (6^th^ November 2020 to 31^st^ January 2021). Note: Multiple tests percentage refers to percentage of the Liverpool population. Ethnicity estimates are following imputation.**

| Measure |  | Uptake (persons) |  | Multiple tests (persons) |  | Positivity (tests) |  |
| --- | --- | --- | --- | --- | --- | --- | --- |
|  |  | Frequency | Percentage | Frequency | Percentage | Frequency | Percentage |
| Total |  | 214525 | 43.1 | 89047 | 17.9 | 5557 | 1.31 |
| Sex | Female | 114517 | 45.9 | 49763 | 20.0 | 2804 | 1.19 |
|  | Male | 100008 | 40.2 | 39284 | 15.8 | 2753 | 1.45 |
| Age band | 0-14 | 19491 | 23.6 | 9029 | 11.0 | 304 | 0.75 |
|  | 15-34 | 78418 | 46.5 | 31685 | 18.8 | 2534 | 1.68 |
|  | 35-69 | 96721 | 49.5 | 41067 | 21.0 | 2498 | 1.25 |
|  | 70+ | 19895 | 38.5 | 7266 | 14.0 | 221 | 0.63 |
| Ethnic group | Asian | 7279 | 37.5 | 2433 | 12.5 | 164 | 1.30 |
|  | Black | 4899 | 39.8 | 1743 | 14.2 | 174 | 2.01 |
|  | Mixed | 3216 | 27.4 | 1349 | 11.5 | 75 | 1.19 |
|  | Other | 2279 | 27.5 | 704 | 8.5 | 113 | 3.11 |
|  | White | 196852 | 47.5 | 82818 | 20.0 | 5031 | 1.27 |
| Deprivation:<br>Liverpool<br>quintiles | Least Deprived | 51957 | 53.0 | 23924 | 24.4 | 1179 | 1.05 |
|  | Quintile 2 | 51625 | 49.1 | 22321 | 21.2 | 1132 | 1.08 |
|  | Quintile 3 | 44248 | 47.0 | 18023 | 19.1 | 1174 | 1.38 |
|  | Quintile 4 | 34679 | 34.5 | 13283 | 13.2 | 1061 | 1.61 |
|  | Most Deprived | 32016 | 31.9 | 11496 | 11.5 | 1011 | 1.75 |
| Deprivation:<br>England<br>quintiles | Least Deprived | 3942 | 58.0 | 2030 | 29.9 | 59 | 0.66 |
|  | Quintile 2 | 27359 | 56.6 | 12922 | 26.7 | 649 | 1.08 |
|  | Quintile 3 | 25832 | 48.6 | 11201 | 21.1 | 607 | 1.13 |
|  | Quintile 4 | 38560 | 47.9 | 16669 | 20.7 | 817 | 1.04 |
|  | Most Deprived | 118832 | 38.4 | 46225 | 15.0 | 3425 | 1.53 |

Trends in the number of tests over time (Figure 1) reflect initial high uptake during the initial push, declining following planned withdrawal of military assistance shortly after Liverpool’s move into less stringent (Tier 2) local restrictions (announced 26^th^ November 2020, enacted 2^nd^ December 2020). Uptake remained initially low in December, before a sharp increase in the week before Christmas as individuals may have sought tests before mixing among Christmas bubbles. High demand was sustained after Christmas and into the national lockdown.

**Figure 1:**
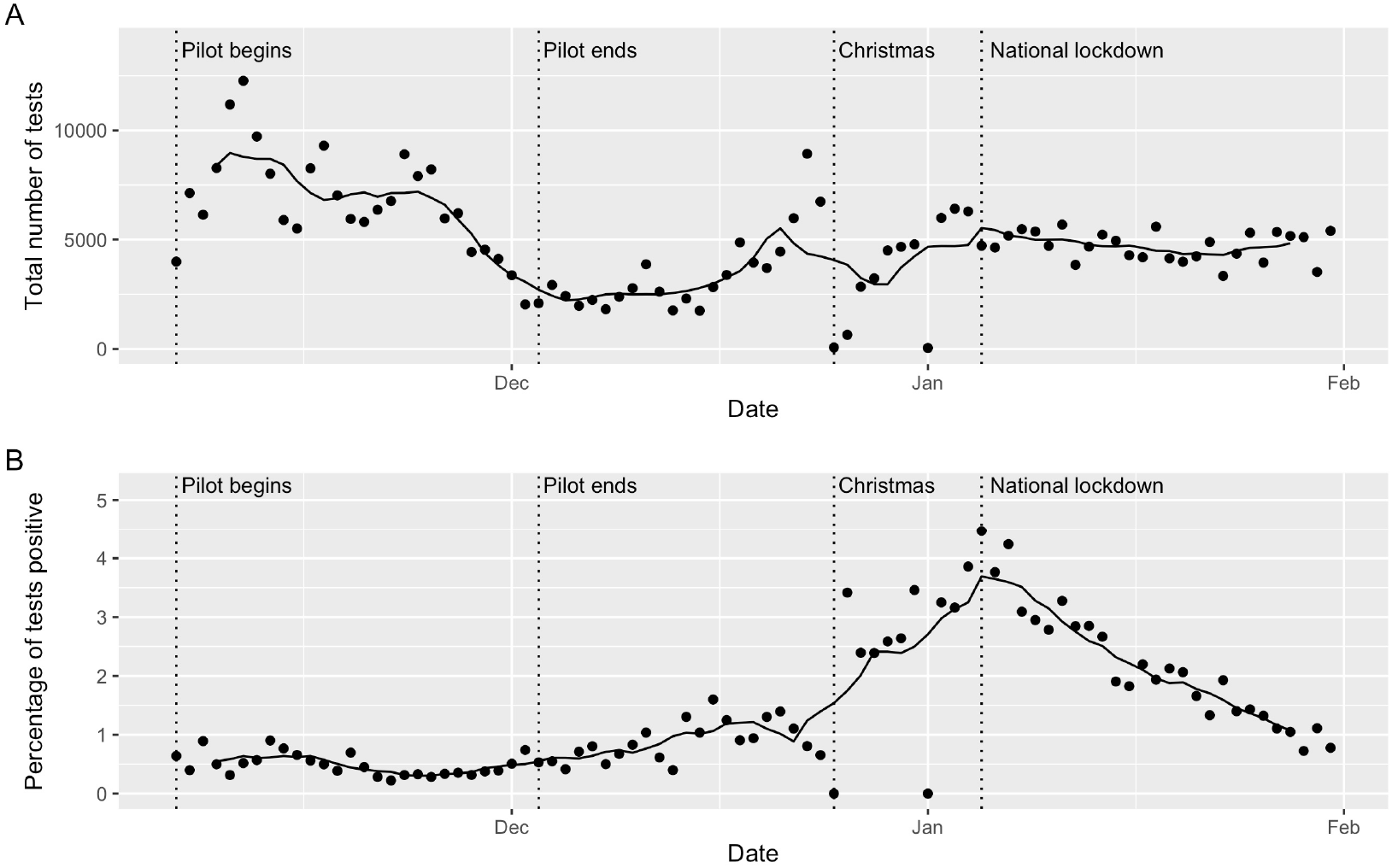
**Trends in the number of lateral flow tests per day (top) and the percentage of lateral flow tests that were positive (bottom). Note: Points are daily values, line is the 7-day average.**

Trends in the positivity rate for LFT remained consistently low (<1.5%) up to Christmas (Figure 1). Post-Christmas there was a rapid increase in the percentage of LFT that were positive, with a doubling of the positivity rate and a small spike around New Year (9-12 days from Christmas). These trends coincided with national increases in COVID-19 being driven (at least in part) by a new variant of the virus. Symptomatic or pauci-symptomatic individuals may have also been accessing asymptomatic testing services during this period due to easier access, quicker turnaround times for test results and habitual changes to testing behaviour, including repeated testing. We examined this hypothesis through exploring trends in individuals accessing LFTs who reported that they had symptoms. A small increase in trends was observed after Christmas (Appendix Figure B2), although overall prevalence remained low (n= 1640 or 0.39% of all LFTs). Positivity rates have declined following the national lockdown.

Figure 2 presents the results from the Bayesian Hierarchical Poisson model exploring the neighbourhood determinants of overall uptake patterns (see Appendix C for full models). Deprivation was negatively related to uptake, suggesting that increasing levels of deprivation were associated with lower uptake. For example, a one standard deviation increase in deprivation score (equivalent of going from Liverpool’s third quintile to most deprived quintile) was associated to 14% fewer tests over the whole period (Relative Risk (RR) = 0.86, 95% Credible Intervals (CIs) = 0.80-0.91). The association was found for each period suggesting the importance of social inequalities in uptake. Distance from home to test site was also important, being negatively associated to uptake suggesting that uptake was lower among those living further from test sites (e.g. whole period RR = 0.95, 95% CIs = 0.91-0.98). Estimating the unstandardized effect size (standardised coefficient / standard deviation) to aid interpretation suggests that each 1km increase in distance to nearest test site was associated with 11% fewer tests. Estimated effect size was largest during the pilot (‘mass testing’) period. While there were also a greater number of test sites during the pilot, the initial choice of sites had been driven by convenience for the Local Authority and military operators, and did not accommodate community perceptions of space, accessability and risks. There was a negative association between the proportion of students in an area and uptake, with effect sizes largest for the two periods post-pilot reflecting that student populations were encouraged to return home in early December (e.g. 6^th^ Jan – 31^st^ Jan RR = 0.91, 95% CIs = 0.87-0.94). Areas that contained a care home were positively associated with uptake, suggesting that testing was higher in areas with a care home present. For example over the whole period, areas with care homes had 15% more tests (RR = 1.15, 95% CIs = 1.07-1.24).

**Figure 2:**
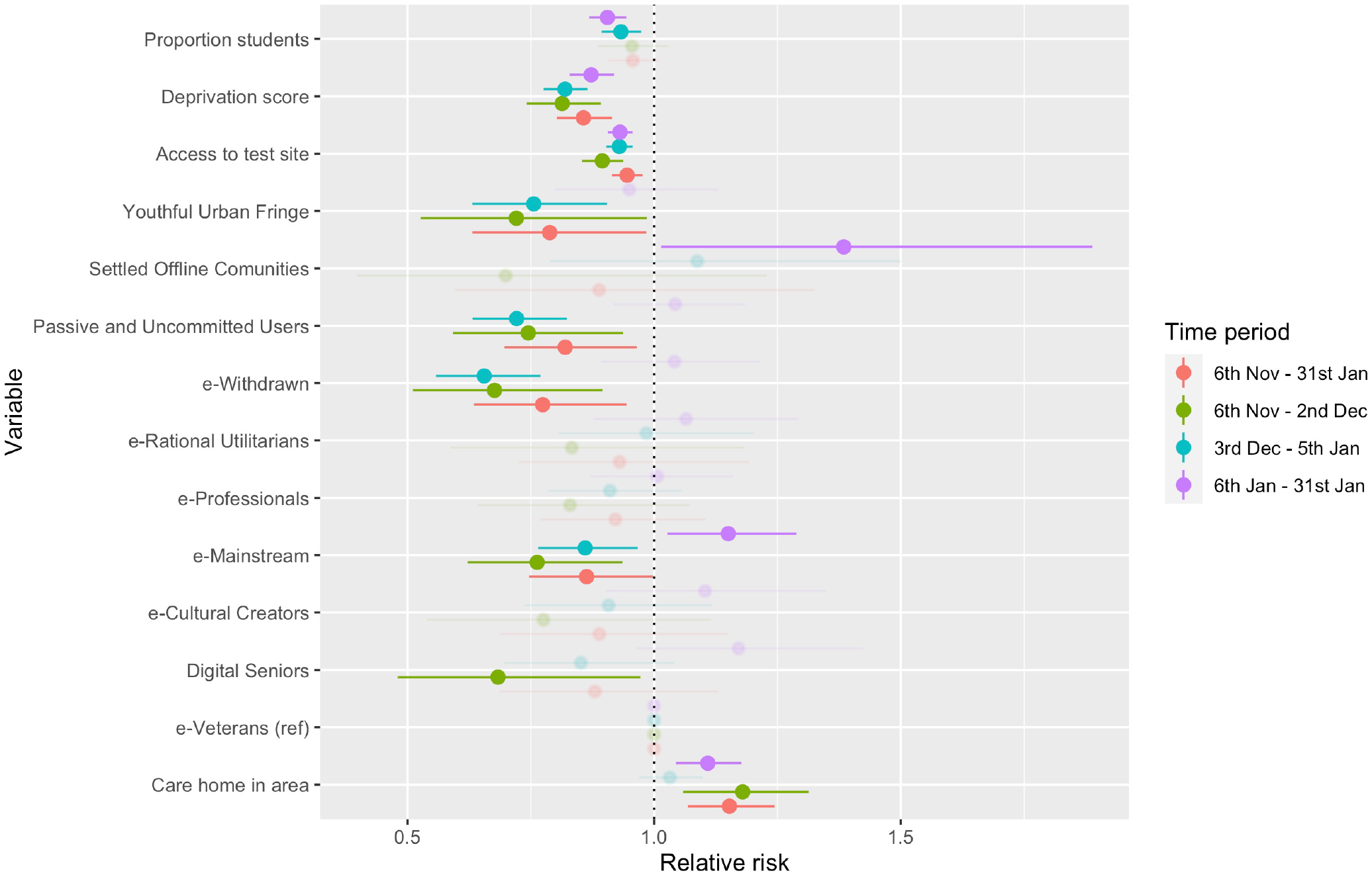
**Estimated relative risks (mean and 95% credible intervals) for the associations between independent variables and uptake of tests by time period model. Note: Transparent values represent estimates where credible intervals contain 1.**

We found the Internet-related characteristics of areas were associated with uptake, suggesting that digital exclusion was a legitimate concern. Populations less confident with using Internet technologies, as measured by the Internet User Classification, showed lower uptake. For example, areas classified as ‘e-Withdrawn’ (described as least engaged with the Internet) had 23% (RR = 0.77, 95% CIs = 0.63-0.94) lower uptake over the whole period than ‘e-Veterans’ (the group hypothesised to have the most confidence with using Internet technologies). Results were inconsistent when using an alternative measure of Internet use (Appendix D).

Analysis for individuals who had multiple LFTs showed similar results to those described above for overall uptake (Figure 3, Appendix C).

**Figure 3:**
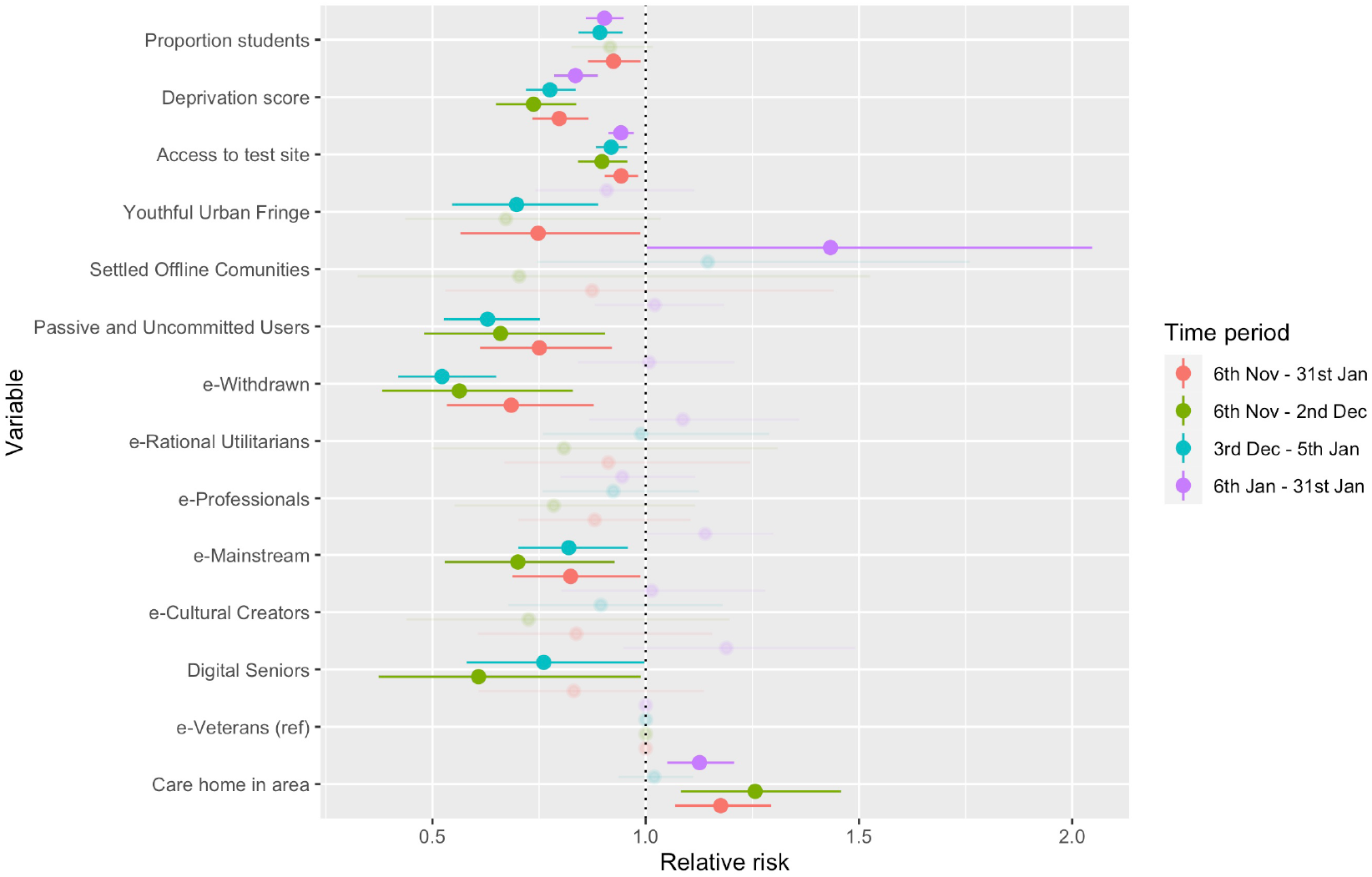
**Estimated relative risks (mean and 95% credible intervals) for the associations between independent variables and multiple tests by time period model. Note: Transparent values represent estimates where credible intervals contain 1.**

Figure 4 presents the model results for positive tests. There was large uncertainty in associations for location of a care home in an area. Deprivation score was positively to positivity at each time period, suggesting that areas that were more deprived had higher proportion of positive tests. For each one standard deviation increase in deprivation score, there was an increase in positive tests by 18% (RR = 1.18, 95% CIs = 1.13-1.24). The proportion of students in an area was negatively associated to positivity for most time periods, with associations uncertain during the initial pilot period. The result suggest that a one standard deviation increase in the proportion of students in an area was associated with 12% fewer positive tests over the whole period (RR = 0.88, 95% CIs = 0.84-0.92). Estimating the unstandardized effect size here to aid interpretation would suggest that a one unit increase in the proportion of students (equivalent to comparing an area where all residents are students to those with none) would see 53% fewer positive tests.

**Figure 4:**
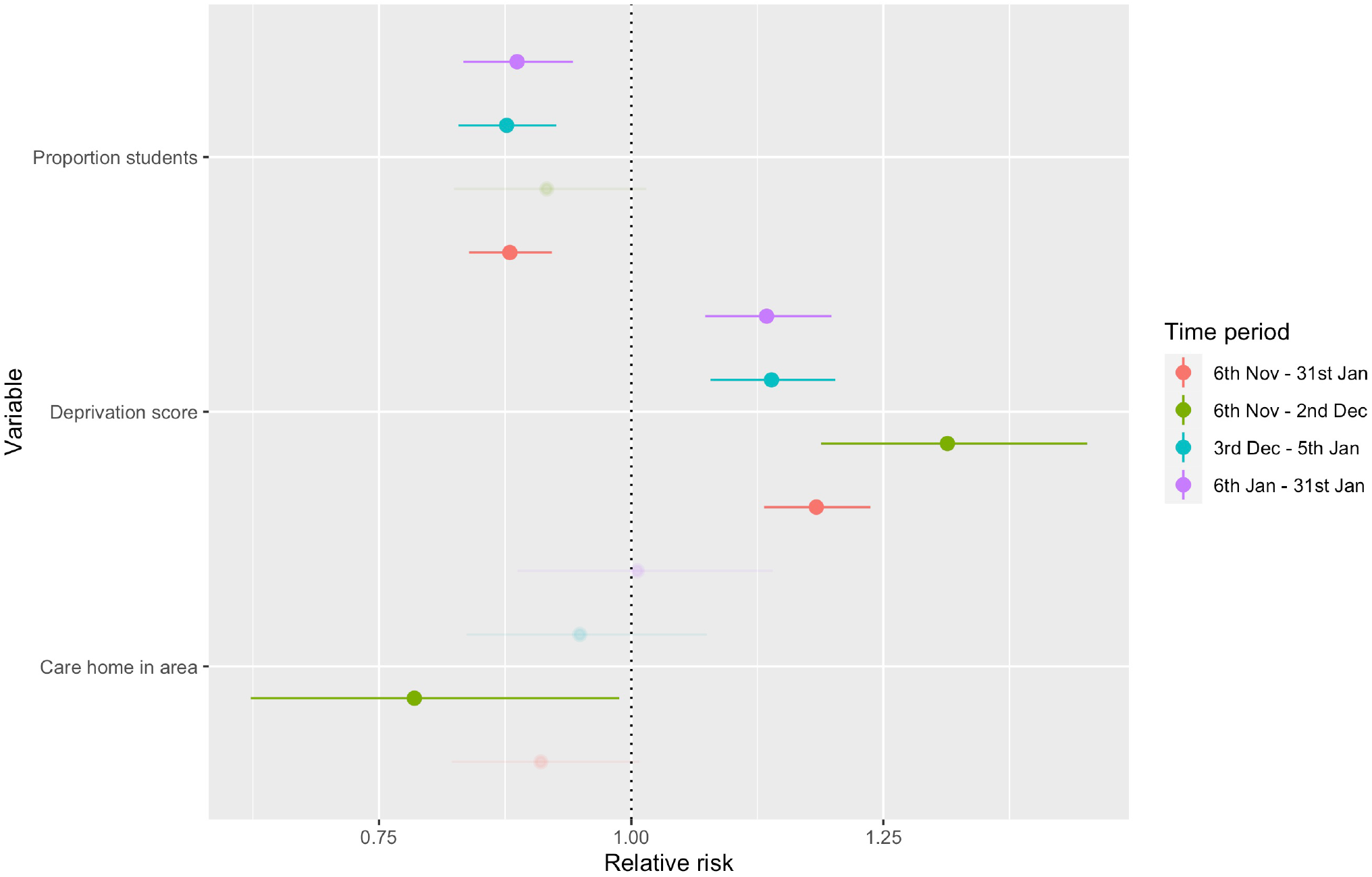
**Estimated relative risks (mean and 95% credible intervals) for the associations between independent variables and positivity by time period model. Note: Transparent values represent estimates where credible intervals contain 1.**

Sensitivity analyses investigating the likelihood of having multiple tests or a positive test at the individual level revealed largely similar associations for the contextual factors described previously (Appendix E). Analyses revealed inequalities by demographic characteristics. Age was negatively associated with the likelihood of a positive test, suggesting that asymptomatic older adults were less likely to have tested positive for COVID-19. Males, compared to females, were more likely to have a positive test and less likely to have had multiple tests. Finally, the ‘Other’ ethnic group were more likely to have had a positive test, with all BAME groups other than ‘Mixed’ less likely to have had multiple tests.

Figures 5 to 7 plot the geographical patterns of the outcome variables estimated from our analytical models. There were distinct geographical inequalities in uptake (similar for both overall uptake and multiple tests), often following patterns of material deprivation with clustering of low uptake in densely populated deprived communities. Geographical patterns were less distinct during the national lockdown, especially for multiple tests, reflecting messages around ‘test if you need to go out to work’ and deprived communities being less likely to be able to work from home. The geographical patterns for uptake contrasted to those for positive tests (Figure 7), which were inversely clustered with higher positivity in deprived areas suggesting spatial inequalities were important in explaining the spread of asymptomatic COVID-19 cases. The contrast between uptake and positive tests is typified through examining the correlation between overall uptake and positivity relative risks, which was −0.54 for the whole study period. The patterns suggest a disconnect between the populations coming forward for testing and those at greatest risk of being infected.

**Figure 5:**
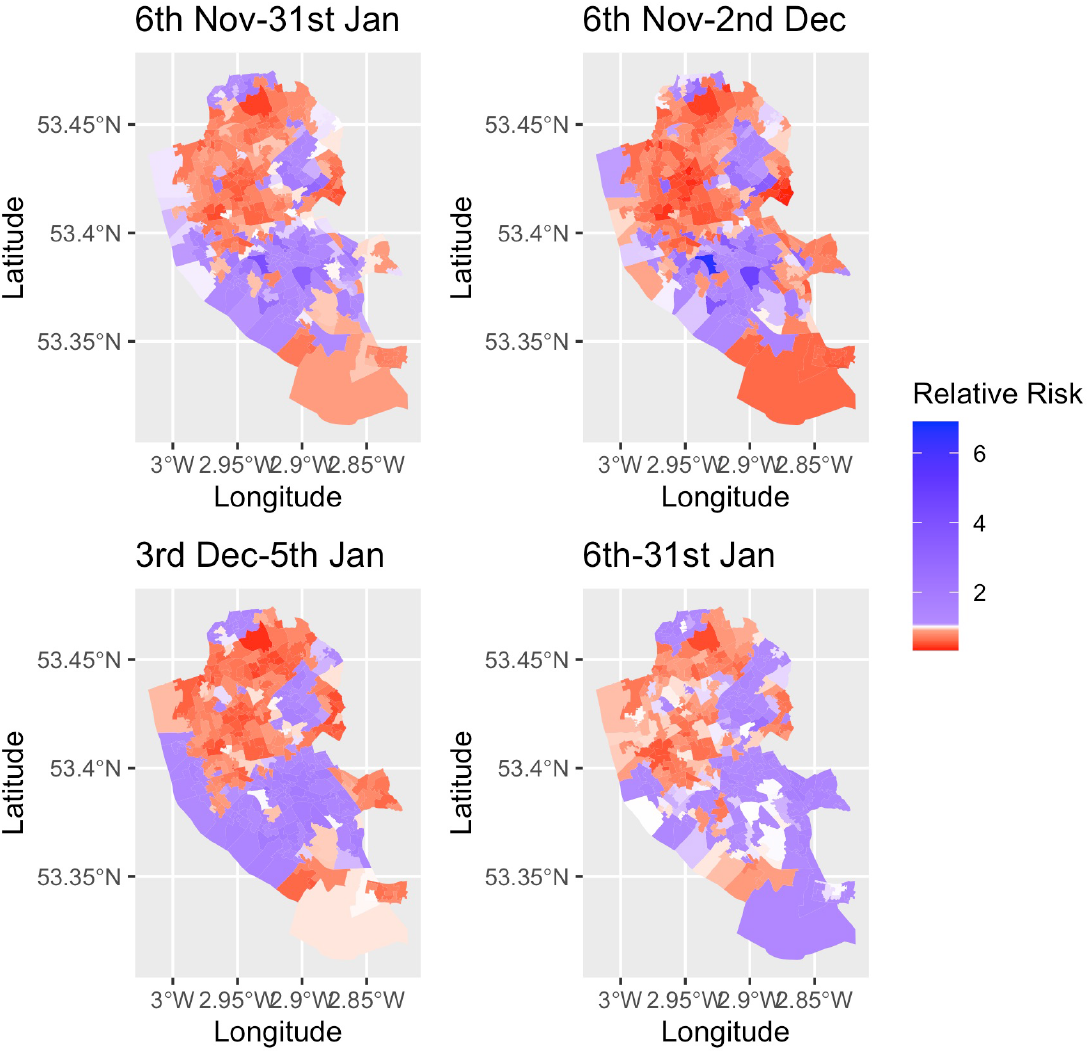
**Relative uptake (observed count / expected count) for overall lateral flow test uptake for lower layer super output areas. Note: red values are relative risks <1, blue colours are >1.**

**Figure 6:**
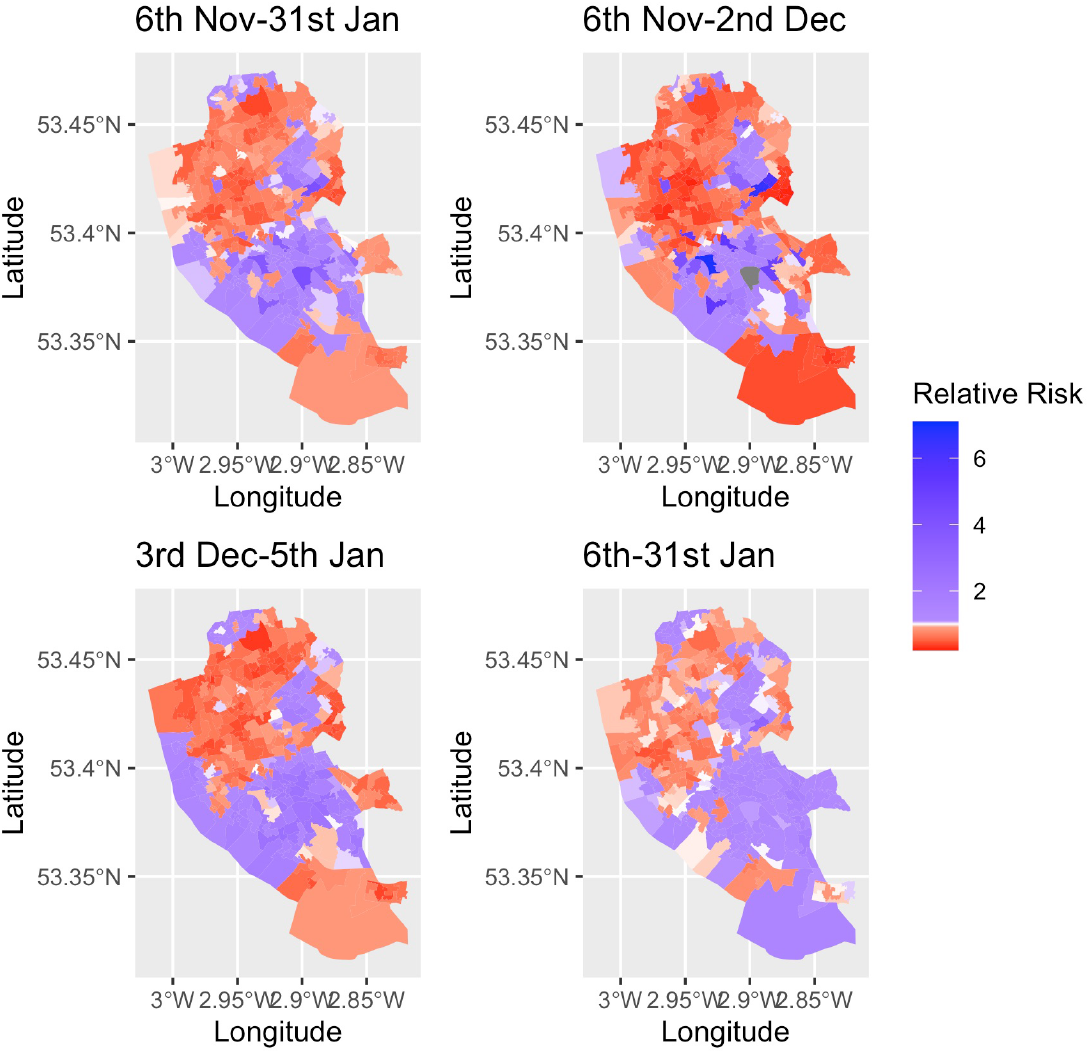
**Relative uptake (observed count / expected count) for multiple lateral flow tests for lower super output areas. Note: red values are relative risks <1, blue colours are >1.**

**Figure 7:**
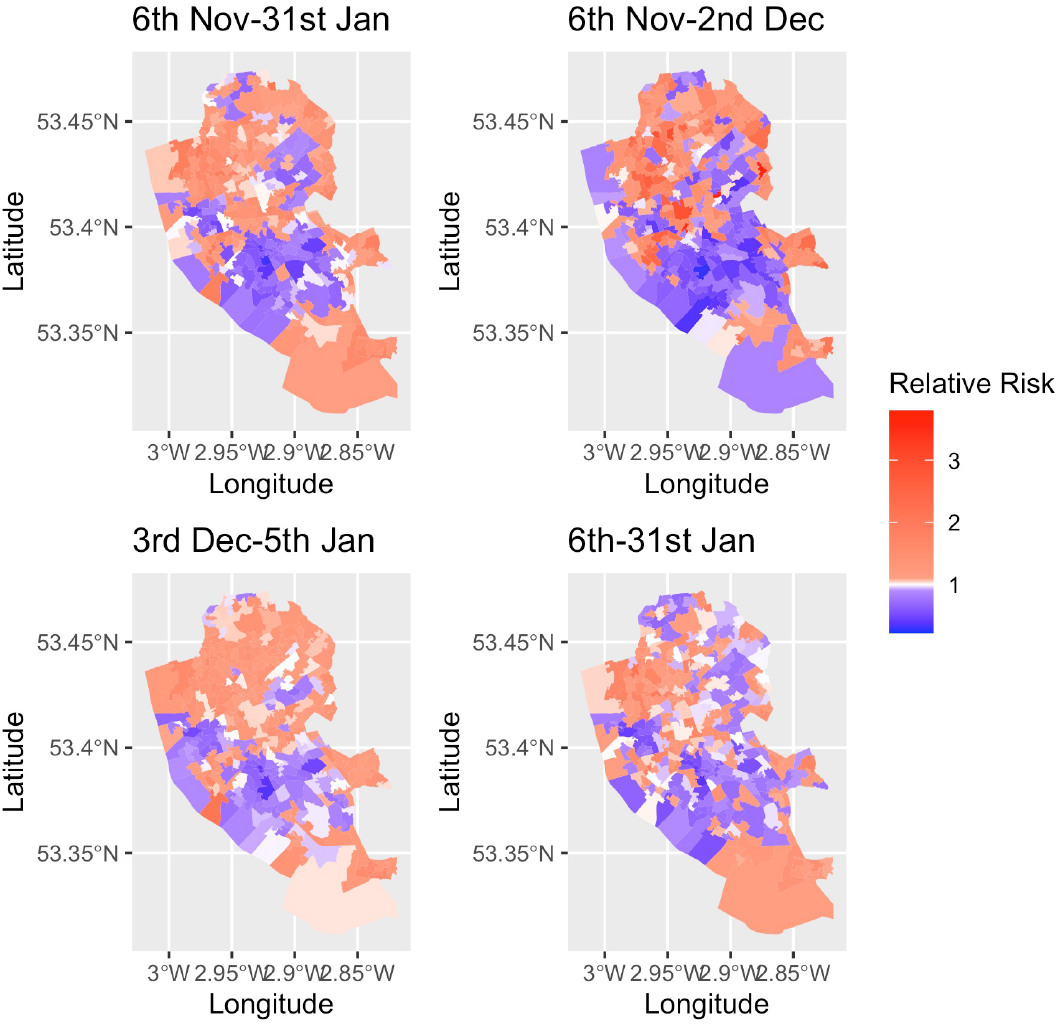
**Relative rates (observed count / expected count) for positive lateral flow tests for lower super output areas. Note: red values are relative risks >1, blue colours are <1.**

## Discussion

Our study provides the first substantial evidence on inequalities involved in large-scale asymptomatic rapid testing of populations for SARS-CoV-2. We find that the provision of free asymptomatic testing was popular with 43% (n = 214 525) residents receiving tests between 6^th^ November 2020 and 31^st^ January 2021. Test positivity (1.3%) was consistent with estimated asymptomatic population prevalence, and 5557 individuals who did not know they had the virus were quickly (within an hour) notified of the need to self-isolate, potentially breaking chains of transmission. Supply and demand for asymptomatic testing was highest during the initial ‘mass testing’ period with military assistance but rose again as SMART testing was introduced with a smaller number of testing centres. Demand was particularly high in the pre-Christmas period, and sustained unexpectedly through lockdown as the advertising message shifted to testing front-line workers. We found evidence of inequalities in uptake and repeat testing, with lower uptake among deprived populations, BAME populations, areas with poor access to test sites and areas classified with high digital exclusion. Spatial inequalities were key in explaining positivity rates, with some evidence of higher positivity among deprived populations and those with low student populations.

There are strengths and weaknesses to our study. We use timely data covering all tests within Liverpool to promptly evaluate a key COVID-19 policy area with little prior evidence. Data were linked to novel geospatial information to contextualise patterns in uptake. Whilst the geospatial data were valuable, there were some discrepancies in data coverage and timing. Although neighbourhood characteristics tend to occur on longer-term trends rather than annual fluctuations [16], our analysis highlights the difficulty in the need for timely socio-economic data for making informed decisions. Our models are cross-sectional and association-based, limiting any causal interpretation. Observations are area based and thus susceptible to ecologic fallacy, which we have attempted to mitigate in our interpretations (also see Appendix E). Analyses are undertaken for small statistical zones that may not reflect actual neighbourhoods, and their defined shapes and sizes may influence the results.

Our study shows that provision of free and voluntary asymptomatic community testing is affected by substantial social and spatial inequalities, typical of the ‘inverse care’ law but with a distinctive digital exclusion factor consistent with the digitally intensive means of accessing testing (participants are usually registered via smartphone and receive results by text message or email, with work-arounds for those without mobile phones). We found large relative inequalities by level of deprivation in uptake, repeat testing and positivity rates. Although uptake was lowest in the most deprived areas, we find that it was higher than the 4% figure shared by others and note no single LSOA had such low uptake [17]. We further identify inequalities by ethnicity and geographical location. The experiences described in our study follow a large body of evidence demonstrating how voluntary or downstream interventions that rely on individual agency often widen the inequalities they seek to tackle [6,7]. These issues are paramount given that the groups we describe as having lower uptake are the groups hit hardest by COVID-19 prevalence and related health and social outcomes [9,10]. Our results suggest that those populations which have lowest uptake tend to be those who likely need it the most.

One year into COVID-19 and societies are still learning how to manage this pandemic. Asymptomatic transmission is a major risk to manage [2], but there is little evidence on how to do so effectively and equitably with rapid tests of infectiousness such as LFT. Our study adds critical and timely evidence. With national expansions planned for the UK and USA, successful management will need to proactively account for the inequalities we describe. Digital inclusion will be key, through engaging with populations less confident in Internet technologies and offering non-digital routes for testing embedded in deprived communities. Test sites will need to be geographically accessible and account for a lack of private transport which is often more common among deprived populations [18]. Emerging evidence suggests that individuals from low income backgrounds may avoid testing, not engage with contact tracing or not isolate if it meant not being able to work [10,19,20]. Greater financial support for individuals isolating may be effective here. Avoiding inequalities in COVID-19 related outcomes is possible through carefully designed interventions, especially when combined into a comprehensive set of interventions.

Future work should identify if large scale asymptomatic community testing can (cost-effectively) break chains of transmission to reduce inequalities. Further work is under way to evaluate whether Liverpool’s community testing pilot impacted case-rates and hospitalisation rates, including if impacts were greater among deprived or vulnerable populations. Identifying barriers to uptake within an inequalities framework – including research explaining the mechanisms that underly the patterns we report – will be important for future policies. Better communication of test accuracy versus purpose is also needed to ensure public confidence and understanding of how to use tests and act on their results [20–22]. The biological, behavioural and operational/systems aspects of testing cannot be separated, and inequalities operate across all these dimensions. Needs-focused and adaptive testing and vaccination policies might be developed to reduce the inequalities that this pandemic is widening.

## Data Availability

Data are accessible via CIPHA. Requests can be made to the Data Access Committee for extracts of the larger-scale data which cannot be released openly due to information governance requirements. All R code is accessible here https://github.com/markagreen/asymptomatic_testing_evaluation.

## Acknowledgements

We thank all the Liverpool residents who participated in at least one test during the study period. Thank you to the team behind CIPHA, including Liverpool Clinical Commissioning group, for their support in generating the data systems behind our analysis. We would like to thank Alessia Calafiore and Alex Singleton for their advice with generating accessibility metrics.

## Contributors

MAG, IB, MGF and BB designed the study. MA, SS and IB were involved in the design, roll-out and delivery of asymptomatic testing in Liverpool. GB, DH and CC set up data systems and supported data cleaning. MAG and BB further cleaned data for analysis. MAG undertook analyses under the supervision of MGF and BB. MAG, IB, BB, MGF, GB, DH and CC provided interpretation for data. All authors contributed to writing the manuscript and agreed to submit the manuscript for publication.

## Ethical approval

The University of Liverpool has provided secondary data analysis as part of a national service evaluation with data collected by Department of Health and Social Care (Sponsor) for quality assurance of Innova lateral flow tests in a public health service intervention. There was no research commissioned by Department of Health and Social Care on this aspect of the Liverpool pilot of asymptomatic, community testing. As such, research ethics approval was not sought by the Department of Health and Social Care. Some aspects of the evaluation requiring fieldwork and primary data collection by the University of Liverpool were subject to ethical approval, which was confirmed prior to the commencement of activities by the University of Liverpool’s Research Ethics Committee. The provision of secondary data analysis and interpretation did not require further ethical approval. Cheshire & Merseyside Health & Care Partnership Combined Intelligence for Population Health Action (CIPHA) Data Access Committee approved access to the data and analysis contained in the study. MAST/SMART was defined as ‘an emergency public health intervention during an extraordinary event’ which were subject to the legal and ethical provisions of a health protection activity and COVID-19 specifically. The secondary analysis of data provided in a health protection activity is not classified as research, and so does not require research ethics committee review (see http://www.hra-decisiontools.org.uk/research/docs/DefiningResearchTable_Oct2017-1.pdf).

## Transparency

The lead author (MAG) affirms that the manuscript is an honest, accurate, and transparent account of the study being reported; that no important aspects of the study have been omitted; and that any discrepancies from the study as planned have been explained.

## References

1 Wei W, Li Z, Chiew C, et al. Presymptomatic Transmission of SARS-CoV-2 — Singapore, January 23–March 16, 2020. Morb Mortal Wkly Rep 2020;69:411–5. doi:http://dx.doi.org/10.15585/mmwr.mm6914e1externalicon

2 Johansson MA, Quandelacy TM, Kada S, et al. SARS-CoV-2 Transmission From People Without COVID-19 Symptoms. JAMA Netw Open 2021;4:e2035057–e2035057. doi:10.1001/jamanetworkopen.2020.35057

3 Buitrago-Garcia D, Egli-Gany D, Counotte MJ, et al. Occurrence and transmission potential of asymptomatic and presymptomatic SARS-CoV-2 infections: A living systematic review and meta-analysis. PLOS Med 2020;17:e1003346. https://doi.org/10.1371/journal.pmed.1003346

4 Department of Health and Social Care. Liverpool to be regularly tested for coronavirus in first whole city testing pilot. 2020. https://www.gov.uk/government/news/liverpool-to-be-regularly-tested-for-coronavirus-in-first-whole-city-testing-pilot

5 Iacobucci G. Covid-19: Government ramps up ‘Moonshot’ mass testing. BMJ 2020;371. doi:10.1136/bmj.m4460

6 McGill R, Anwar E, Orton L, et al. Are interventions to promote healthy eating equally effective for all? Systematic review of socioeconomic inequalities in impact. BMC Public Health 2015;15:457. doi:10.1186/s12889-015-1781-7

7 McLaren L, McIntyre L, Kirkpatrick S. Rose’s population strategy of prevention need not increase social inequalities in health. Int J Epidemiol 2010;39:372–7. doi:10.1093/ije/dyp315

8 Dodds C, Mugweni E, Phillips G, et al. Acceptability of HIV self-sampling kits (TINY vial) among people of black African ethnicity in the UK: a qualitative study. BMC Public Health 2018;18:499. doi:10.1186/s12889-018-5256-5

9 Public Health England. Disparities in the risk and outcomes of COVID-19. 2020. https://assets.publishing.service.gov.uk/government/uploads/system/uploads/attachment_data/file/908434/Disparities_in_the_risk_and_outcomes_of_COVID_Aug ust_2020_update.pdf

10 Paremoer L, Nandi S, Serag H, et al. Covid-19 pandemic and the social determinants of health. BMJ 2021;372:129.

11 Office for National Statistics. Population estimates. 2020. https://www.ons.gov.uk/peoplepopulationandcommunity/populationandmigration/populationestimates

12 Ministry of Housing Communities & Local Government. English indices of deprivation 2019. 2020. https://www.gov.uk/government/statistics/english-indices-of-deprivation-2019

13 Singleton A, Alexiou A, Savani R. Mapping the geodemographics of digital inequality in Great Britain: An integration of machine learning into small area estimation. Comput Environ Urban Syst 2020;82:101486. doi:https://doi.org/10.1016/j.compenvurbsys.2020.101486

14 Besag J, York J, Mollié A. Bayesian image restoration, with two applications in spatial statistics. Ann Inst Stat Math 1991;43:1–20. doi:10.1007/BF00116466

15 Rue H, Riebler A, Sørbye SH, et al. Bayesian Computing with INLA: A Review. Annu Rev Stat Its Appl 2017;4:395–421. doi:10.1146/annurev-statistics-060116-054045

16 Singleton A, Pavlis M, Longley PA. The stability of geodemographic cluster assignments over an intercensal period. J Geogr Syst 2016;18:97–123. doi:10.1007/s10109-016-0226-x

17 Wise J. Covid-19: Concerns persist about purpose, ethics, and effect of rapid testing in Liverpool. BMJ 2020;371:m4690.

18 Lucas K, Stokes G, Bastiaanssen J, et al. Inequalities in Mobility and Access in the UK Transport System. 2019. https://assets.publishing.service.gov.uk/government/uploads/system/uploads/attachment_data/file/784685/future_of_mobility_access.pdf

19 Briggs ADM, Fraser C. Is NHS Test and Trace exacerbating COVID-19 inequalities? Lancet 2020;396:1972. doi:10.1016/S0140-6736(20)32593-9

20 University of Liverpool. Liverpool Covid-19 Community Testing Pilot - Interim Evaluation Report. 2020. https://www.liverpool.ac.uk/coronavirus/research-and-analysis/covid-smart-pilot/

21 Mina MJ, Parker R, Larremore DB. Rethinking Covid-19 Test Sensitivity — A Strategy for Containment. N Engl J Med 2020;383:e120. doi:10.1056/NEJMp2025631

22 Crozier A, Rajan S, Buchan I, et al. Put to the test: use of rapid testing technologies for covid-19. BMJ 2021;372:208.

