## Supplementary Appendices for "Evaluating social and spatial inequalities of large scale rapid lateral flow SARS-CoV-2 antigen testing in COVID-19 management: An observational study of Liverpool, UK (November 2020 to January 2021)"

The document presents additional data and analyses to support the results presented in the manuscript entitled '*Evaluating social and spatial inequalities of large scale rapid lateral flow SARS-CoV-2 antigen testing in COVID-19 management: An observational study of Liverpool, UK (November 2020 to January 2021)*'.

#### Appendix A: Missing data

From the CIPHA database containing 1 007 723 lateral flow tests between 6th November 2020 and 31st January 2021, we identified 425 593 lateral flow tests for residents in Liverpool (5955 tests or 0.006% had missing address data). Investigating patterns in missing data for our analytical dataset (i.e., the 425 593 lateral flow tests) revealed no missing data for sex. All tests contained information about age, however we treated all tests with age less than or equal to 5 years as missing (1.6%) since there was no asymptomatic tests being delivered to these ages. The high level of data completion was captured via users completing electronic forms on site prior to their test (recommended on their mobile phones), with additional information gathered from data linkage to NHS data systems. Where records for individuals had different recorded ages (i.e. individuals who had birthdays over the study period), we selected the earliest test. Tests with void or insufficient results (n=1755, 0.4%) were excluded from analyses.

Ethnicity had lower completed coverage with 9.8% (n = 41 640) tests with missing data post-data linkage, with individuals selecting 'prefer not to say' when registering at a test centre. We used imputation by polytomous regression to impute ethnic groups for persons with missing data using Multivariate Imputation by Chained Equations (R packaged 'mice'). The method is commonly used for imputing missing data. The approach assumes that records were missing at random and this assumption may not hold with our data. Predicted ethnic group (White, Black, Asian, Mixed or Other) was estimated based on an individual's age, sex and the composition of their neighbourhood (lower super output area). Area level predictors included the proportion of residents by each ethnic group, deprivation score, the proportion of students, and the number of care home beds per population.

**Table A1: Frequency of missing data by test records for Liverpool.**

| Variable | Frequency | Percentage |
|---|---|---|
| Age | 6821 | 1.6% |
| Sex | 0 | 0% |
| Ethnicity | 41640 | 9.8% |
| Test result* | 1755 | 0.4% |

* Refers to tests with void or insufficient results

#### Appendix B: Descriptive statistics

Detailed descriptive statistics for each outcome measure by time period of analysis are presented in Table B1-B3.

**Table B1: Summary statistics for individuals who received at least one lateral flow antigen test by population characteristics.**

| Measure | | 6 Nov 2020 - 13 Jan 2021 | | 6 Nov - 2 Dec 2020 | | 3 Dec 2020 - 5 Jan 2021 | | 6 Jan - 31 Jan 2021 | |
|---|---|---|---|---|---|---|---|---|---|
| | | Frequency | Percentage | Frequency | Percentage | Frequency | Percentage | Frequency | Percentage |
| Total persons | | 214525 | 43.1 | 117549 | 23.6 | 80981 | 16.3 | 75371 | 15.1 |
| Sex | Female | 114517 | 45.9 | 63991 | 25.7 | 43929 | 17.6 | 41253 | 16.5 |
| | Male | 100008 | 40.2 | 53558 | 21.5 | 37052 | 14.9 | 34118 | 13.7 |
| Age band | 0-14 | 19491 | 23.6 | 14905 | 18.1 | 4350 | 5.3 | 3029 | 3.7 |
| | 15-34 | 78418 | 46.5 | 35803 | 21.2 | 32597 | 19.3 | 28366 | 16.8 |
| | 35-69 | 96721 | 49.5 | 52433 | 26.8 | 37776 | 19.3 | 39461 | 20.2 |
| | 70+ | 19895 | 38.5 | 14408 | 27.8 | 6258 | 12.1 | 4515 | 8.7 |
| Ethnic group | Asian | 7279 | 37.5 | 4026 | 20.7 | 2221 | 11.4 | 2196 | 11.3 |
| | Black | 4899 | 39.8 | 2437 | 19.8 | 1533 | 12.5 | 1819 | 14.8 |
| | Mixed | 3216 | 27.4 | 1702 | 14.5 | 1225 | 10.4 | 1072 | 9.1 |
| | Other | 2279 | 27.5 | 1078 | 13.0 | 752 | 9.1 | 787 | 9.5 |
| | White | 196852 | 47.5 | 108306 | 26.1 | 75250 | 18.1 | 69497 | 16.8 |
| Deprivation: Liverpool quintiles | Least Deprived | 51957 | 53.0 | 29949 | 30.5 | 22866 | 23.3 | 16752 | 17.1 |
| | Quintile 2 | 51625 | 49.1 | 28372 | 27.0 | 20587 | 19.6 | 17309 | 16.5 |
| | Quintile 3 | 44248 | 47.0 | 25112 | 26.7 | 15170 | 16.1 | 15241 | 16.2 |
| | Quintile 4 | 34679 | 34.5 | 18150 | 18.1 | 11611 | 11.6 | 13296 | 13.2 |
| | Most Deprived | 32016 | 31.9 | 15966 | 15.9 | 10747 | 10.7 | 12773 | 12.7 |
| Deprivation: England quintiles | Least Deprived | 3942 | 58.0 | 2583 | 38.0 | 1593 | 23.4 | 964 | 14.2 |
| | Quintile 2 | 27359 | 56.6 | 16316 | 33.7 | 12184 | 25.2 | 8540 | 17.7 |
| | Quintile 3 | 25832 | 48.6 | 13603 | 25.6 | 11492 | 21.6 | 9105 | 17.1 |
| | Quintile 4 | 38560 | 47.9 | 21423 | 26.6 | 15199 | 18.9 | 12793 | 15.9 |
| | Most Deprived | 118832 | 38.4 | 63624 | 20.6 | 40513 | 13.1 | 43969 | 14.2 |

**Table B2: Summary statistics for individuals who received multiple tests by population characteristics.**

| Measure | | 6 Nov 2020 - 13 Jan 2021 | | 6 Nov - 2 Dec 2020 | | 3 Dec 2020 - 5 Jan 2021 | | 6 Jan - 31 Jan 2021 | |
|---|---|---|---|---|---|---|---|---|---|
| | | Frequency | Percentage | Frequency | Percentage | Frequency | Percentage | Frequency | Percentage |
| Total persons | | 89047 | 17.9 | 40776 | 8.2 | 23071 | 4.6 | 24250 | 4.9 |
| Sex | Female | 49763 | 20.0 | 22806 | 9.1 | 12835 | 5.1 | 14125 | 5.7 |
| | Male | 39284 | 15.8 | 17970 | 7.2 | 10236 | 4.1 | 10125 | 4.1 |
| Age band | 0-14 | 9029 | 11.0 | 7356 | 8.9 | 1421 | 1.7 | 1049 | 1.3 |
| | 15-34 | 31685 | 18.8 | 12042 | 7.1 | 9659 | 5.7 | 9209 | 5.5 |
| | 35-69 | 41067 | 21.0 | 16914 | 8.7 | 10574 | 5.4 | 12990 | 6.7 |
| | 70+ | 7266 | 14.0 | 4464 | 8.6 | 1417 | 2.7 | 1002 | 1.9 |
| Ethnic group | Asian | 2433 | 12.5 | 1129 | 5.8 | 508 | 2.6 | 701 | 3.6 |
| | Black | 1743 | 14.2 | 716 | 5.8 | 342 | 2.8 | 588 | 4.8 |
| | Mixed | 1349 | 11.5 | 625 | 5.3 | 341 | 2.9 | 354 | 3.0 |
| | Other | 704 | 8.5 | 275 | 3.3 | 157 | 1.9 | 204 | 2.5 |
| | White | 82818 | 20.0 | 38031 | 9.2 | 21723 | 5.2 | 22403 | 5.4 |
| Deprivation: Liverpool quintiles | Least Deprived | 23924 | 24.4 | 11596 | 11.8 | 7351 | 7.5 | 5741 | 5.9 |
| | Quintile 2 | 22321 | 21.2 | 10590 | 10.1 | 6140 | 5.8 | 5690 | 5.4 |
| | Quintile 3 | 18023 | 19.1 | 8470 | 9.0 | 4163 | 4.4 | 4873 | 5.2 |
| | Quintile 4 | 13283 | 13.2 | 5700 | 5.7 | 2939 | 2.9 | 4109 | 4.1 |
| | Most Deprived | 11496 | 11.5 | 4420 | 4.4 | 2478 | 2.5 | 3837 | 3.8 |
| Deprivation: England quintiles | Least Deprived | 2030 | 29.9 | 1244 | 18.3 | 520 | 7.6 | 363 | 5.3 |
| | Quintile 2 | 12922 | 26.7 | 6466 | 13.4 | 4002 | 8.3 | 2937 | 6.1 |
| | Quintile 3 | 11201 | 21.1 | 4783 | 9.0 | 3580 | 6.7 | 3036 | 5.7 |
| | Quintile 4 | 16669 | 20.7 | 8089 | 10.0 | 4472 | 5.6 | 4240 | 5.3 |
| | Most Deprived | 46225 | 15.0 | 20194 | 6.5 | 10497 | 3.4 | 13674 | 4.4 |

**Table B3: Summary statistics for the numbers of positive tests. Note: Numbers <10 have been redacted due to statistical disclosure issues.**

| Measure | | 6 Nov 2020 - 13 Jan 2021 | | 6 Nov - 2 Dec 2020 | | 3 Dec 2020 - 5 Jan 2021 | | 6 Jan - 31 Jan 2021 | |
|---|---|---|---|---|---|---|---|---|---|
| | | Frequency | Percentage | Frequency | Percentage | Frequency | Percentage | Frequency | Percentage |
| Total tests | | 425793 | | 183276 | | 119939 | | 122578 | |
| Total positive tests | | 5557 | 1.31 | 871 | 0.48 | 2107 | 1.76 | 2579 | 2.10 |
| Sex | Female | 2804 | 1.19 | 463 | 0.46 | 1100 | 1.68 | 1241 | 1.80 |
| | Male | 2753 | 1.45 | 408 | 0.49 | 1007 | 1.85 | 1338 | 2.49 |
| Age band | 6-14 | 304 | 0.75 | 94 | 0.33 | 97 | 1.47 | 113 | 2.16 |
| | 15-34 | 2534 | 1.68 | 348 | 0.62 | 1061 | 2.18 | 1125 | 2.45 |
| | 35-69 | 2498 | 1.25 | 368 | 0.47 | 869 | 1.55 | 1261 | 1.93 |
| | 70+ | 221 | 0.63 | 61 | 0.30 | 80 | 0.94 | 80 | 1.31 |
| Ethnic group | Asian | 164 | 1.30 | 44 | 0.76 | 49 | 1.59 | 71 | 1.93 |
| | Black | 174 | 2.01 | 34 | 0.95 | 45 | 2.15 | 95 | 3.18 |
| | Mixed | 75 | 1.19 | 15 | 0.53 | 27 | 1.54 | 33 | 1.91 |
| | Other | 113 | 3.11 | 22 | 1.46 | 52 | 5.33 | 39 | 3.39 |
| | White | 5031 | 1.27 | 756 | 0.45 | 1934 | 1.73 | 2341 | 2.07 |
| Deprivation: Liverpool quintiles | Least Deprived | 1179 | 1.05 | 157 | 0.32 | 516 | 1.46 | 506 | 1.82 |
| | Quintile 2 | 1132 | 1.08 | 178 | 0.39 | 483 | 1.57 | 471 | 1.64 |
| | Quintile 3 | 1174 | 1.38 | 206 | 0.54 | 438 | 1.98 | 530 | 2.16 |
| | Quintile 4 | 1061 | 1.61 | 166 | 0.60 | 346 | 2.07 | 549 | 2.57 |
| | Most Deprived | 1011 | 1.75 | 164 | 0.72 | 324 | 2.17 | 523 | 2.60 |
| Deprivation: England quintiles | Least Deprived | 59 | 0.66 | <10 | -- | 22 | 0.88 | 31 | 1.84 |
| | Quintile 2 | 649 | 1.08 | 87 | 0.32 | 280 | 1.47 | 282 | 1.99 |
| | Quintile 3 | 607 | 1.13 | 84 | 0.39 | 273 | 1.57 | 250 | 1.66 |
| | Quintile 4 | 817 | 1.04 | 134 | 0.39 | 332 | 1.46 | 351 | 1.64 |
| | Most Deprived | 3425 | 1.53 | 560 | 0.58 | 1200 | 2.06 | 1665 | 2.37 |

Figure B1 presents the estimated percentage of the population in Liverpool who received a lateral flow test by 10 year age band. Uptake was lowest among the 6-9 and 80+ age groups, and highest among the 10-19 age group. The high uptake among individuals aged 10-19 mostly represents targeted testing among University students, as well as pilots targeting secondary schools and colleges.

Figure B2 plots trends in the percentage of lateral flow tests which were for individuals who reported symptoms of COVID-19. Overall prevalence was low (n = 1640, 0.39%). While the temporal trend remains low throughout the period, there are periods where the prevalence was higher including the start of the pilot and immediately following Christmas (the latter period reflecting a doubling of the previous week's values). Individuals could report whether they had any COVID-19 symptoms in the last 72 hours when registering their details prior to taking a lateral flow test. As such, these data are self-reported and likely to have under-estimated the numbers of symptomatic individuals who received lateral flow tests. If an individual showed up for a lateral flow test and said they had symptoms, the protocol was that they should have been redirected to a symptomatic test site, however this might have not always happened.

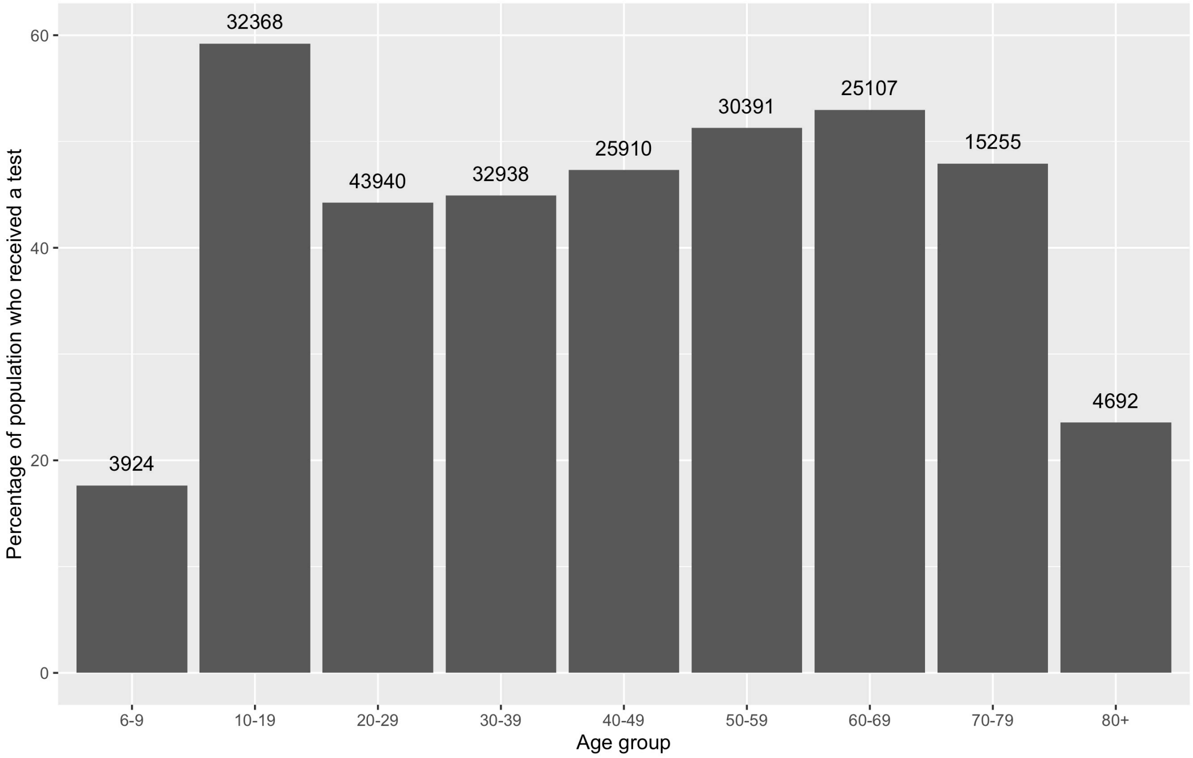

**Figure B1: Percentage of people by ten-year age band who received a lateral flow test between 6th November 2020 and 13th Jan 2021 (frequency counts for uptake plotted above bars).**

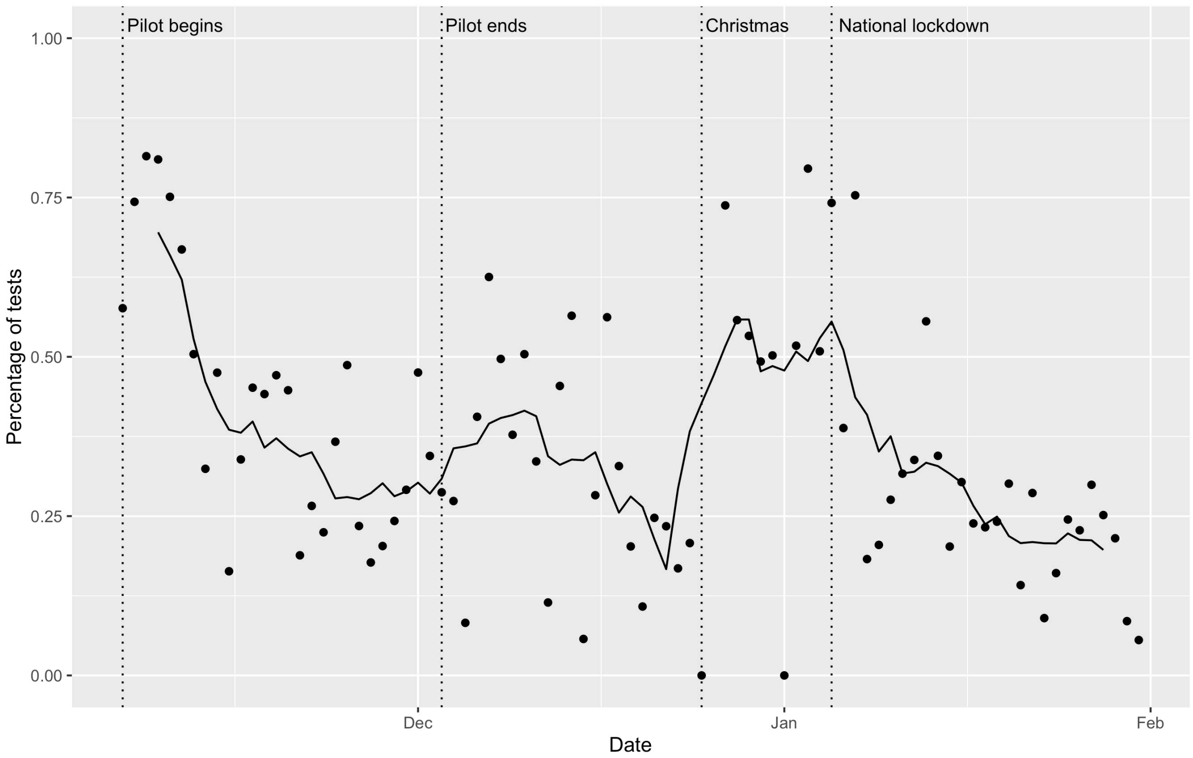

**Figure B2: Percentage of people who received lateral flow tests who reported that they had symptoms. Note: points represent the raw daily value, with the line the 7 day average.**

### Appendix C: Full analytical model results

The section presents full tables of the spatial regression models that were presented in the main paper (Figures 2 to 4).

**Table C1: Relative risks (RR) for the associations between covariates and people having lateral flow tests per area. Note: Lower and upper limits refer to 95% Credible Intervals. Models adjusted for age, sex and ethnicity using indirect standardisation.**

| Variable | 6th Nov - 13th Jan | | | 6th Nov - 2nd Dec | | | 3rd Dec - 5th Jan | | | 6th Jan - 31st Jan | | |
|---|---|---|---|---|---|---|---|---|---|---|---|---|
| | RR | lower | upper | RR | lower | upper | RR | lower | upper | RR | lower | upper |
| Deprivation score | 0.86 | 0.80 | 0.91 | 0.81 | 0.74 | 0.89 | 0.82 | 0.78 | 0.87 | 0.87 | 0.83 | 0.92 |
| Proportion students | 0.96 | 0.91 | 1.01 | 0.95 | 0.89 | 1.03 | 0.93 | 0.89 | 0.97 | 0.91 | 0.87 | 0.94 |
| Care home in area | 1.15 | 1.07 | 1.24 | 1.18 | 1.06 | 1.31 | 1.03 | 0.97 | 1.10 | 1.11 | 1.04 | 1.18 |
| Access to test site (km) | 0.95 | 0.91 | 0.98 | 0.89 | 0.85 | 0.94 | 0.93 | 0.90 | 0.96 | 0.93 | 0.91 | 0.96 |
| IUC ( reference: e-Veterans) | | | | | | | | | | | | |
| Digital Seniors | 0.88 | 0.69 | 1.13 | 0.68 | 0.48 | 0.97 | 0.85 | 0.70 | 1.04 | 1.17 | 0.96 | 1.42 |
| e-Cultural Creators | 0.89 | 0.69 | 1.15 | 0.78 | 0.54 | 1.12 | 0.91 | 0.74 | 1.12 | 1.10 | 0.90 | 1.35 |
| e-Mainstream | 0.86 | 0.75 | 1.00 | 0.76 | 0.62 | 0.94 | 0.86 | 0.77 | 0.97 | 1.15 | 1.03 | 1.29 |
| e-Professionals | 0.92 | 0.77 | 1.10 | 0.83 | 0.64 | 1.07 | 0.91 | 0.79 | 1.06 | 1.01 | 0.87 | 1.16 |
| e-Rational Utilitarians | 0.93 | 0.73 | 1.19 | 0.83 | 0.59 | 1.18 | 0.98 | 0.81 | 1.20 | 1.06 | 0.88 | 1.29 |
| e-Withdrawn | 0.77 | 0.63 | 0.94 | 0.68 | 0.51 | 0.90 | 0.66 | 0.56 | 0.77 | 1.04 | 0.89 | 1.22 |
| Passive and Uncommitted Users | 0.82 | 0.70 | 0.96 | 0.74 | 0.59 | 0.94 | 0.72 | 0.63 | 0.82 | 1.04 | 0.92 | 1.18 |
| Settled Offline Communities | 0.89 | 0.60 | 1.32 | 0.70 | 0.40 | 1.23 | 1.09 | 0.79 | 1.50 | 1.38 | 1.01 | 1.89 |
| Youthful Urban Fringe | 0.79 | 0.63 | 0.98 | 0.72 | 0.53 | 0.99 | 0.76 | 0.63 | 0.90 | 0.95 | 0.80 | 1.13 |

*Model fit (log marginal-likelihood)* – Model for period 6th Nov – 13th Jan = -2015; Model for period 6th Nov – 2nd Dec = -1918; Model for period 3rd Dec – 5th Jan = -1664; Model for period 6th Jan – 31st Jan = -1644.

**Table C2: Relative risks (RR) for the associations between covariates and people who received multiple lateral flow tests per area. Note: Lower and upper limits refer to 95% Credible Intervals. Models adjusted for age, sex and ethnicity using indirect standardisation.**

| Variable | 6th Nov - 13th Jan | | | 6th Nov - 2nd Dec | | | 3rd Dec - 5th Jan | | | 6th Jan - 31st Jan | | |
|---|---|---|---|---|---|---|---|---|---|---|---|---|
| | RR | lower | upper | RR | lower | upper | RR | lower | upper | RR | lower | upper |
| Deprivation score | 0.80 | 0.73 | 0.87 | 0.74 | 0.65 | 0.84 | 0.78 | 0.72 | 0.84 | 0.84 | 0.79 | 0.89 |
| Proportion students | 0.92 | 0.86 | 0.99 | 0.92 | 0.83 | 1.02 | 0.89 | 0.84 | 0.95 | 0.90 | 0.86 | 0.95 |

| | | | | | | | | | | | | |
|---|---|---|---|---|---|---|---|---|---|---|---|---|
| Care home in area | 1.18 | 1.07 | 1.29 | 1.26 | 1.08 | 1.46 | 1.02 | 0.94 | 1.11 | 1.13 | 1.05 | 1.21 |
| Access to test site | 0.94 | 0.90 | 0.98 | 0.90 | 0.84 | 0.96 | 0.92 | 0.88 | 0.96 | 0.94 | 0.91 | 0.97 |
| IUC (reference: e-Veterans) | | | | | | | | | | | | |
| Digital Seniors | 0.83 | 0.61 | 1.14 | 0.61 | 0.37 | 0.99 | 0.76 | 0.58 | 1.00 | 1.19 | 0.95 | 1.49 |
| e-Cultural Creators | 0.84 | 0.61 | 1.16 | 0.73 | 0.44 | 1.20 | 0.89 | 0.68 | 1.18 | 1.01 | 0.80 | 1.28 |
| e-Mainstream | 0.82 | 0.69 | 0.99 | 0.70 | 0.53 | 0.93 | 0.82 | 0.70 | 0.96 | 1.14 | 1.00 | 1.30 |
| e-Professionals | 0.88 | 0.70 | 1.10 | 0.78 | 0.55 | 1.12 | 0.92 | 0.76 | 1.12 | 0.95 | 0.80 | 1.12 |
| e-Rational Utilitarians | 0.91 | 0.67 | 1.25 | 0.81 | 0.50 | 1.31 | 0.99 | 0.76 | 1.29 | 1.09 | 0.87 | 1.36 |
| e-Withdrawn | 0.68 | 0.53 | 0.88 | 0.56 | 0.38 | 0.83 | 0.52 | 0.42 | 0.65 | 1.01 | 0.84 | 1.21 |
| Passive and Uncommitted Users | 0.75 | 0.61 | 0.92 | 0.66 | 0.48 | 0.91 | 0.63 | 0.53 | 0.75 | 1.02 | 0.88 | 1.18 |
| Settled Offline Communities | 0.87 | 0.53 | 1.44 | 0.70 | 0.32 | 1.53 | 1.15 | 0.75 | 1.76 | 1.43 | 1.00 | 2.05 |
| Youthful Urban Fringe | 0.75 | 0.57 | 0.99 | 0.67 | 0.44 | 1.04 | 0.70 | 0.55 | 0.89 | 0.91 | 0.74 | 1.11 |

*Model fit (log marginal-likelihood)* – Model for period 6$^{th}$ Nov – 13$^{th}$ Jan = -1806; Model for period 6$^{th}$ Nov – 2$^{nd}$ Dec = -1658; Model for period 3$^{rd}$ Dec – 5$^{th}$ Jan = -1365; Model for period 6$^{th}$ Jan – 31$^{st}$ Jan = -1349.

**Table C3: Relative risks (RR) for the associations between covariates and positive lateral flow tests per area. Note: Lower and upper limits refer to 95% Credible Intervals. Models adjusted for age, sex and ethnicity using indirect standardisation.**

| Variable | 6th Nov - 13th Jan | | | 6th Nov - 2nd Dec | | | 3rd Dec - 5th Jan | | | 6th Jan - 31st Jan | | |
|---|---|---|---|---|---|---|---|---|---|---|---|---|
| | RR | lower | upper | RR | lower | upper | RR | lower | upper | RR | lower | upper |
| Deprivation score | 1.18 | 1.13 | 1.24 | 1.31 | 1.19 | 1.45 | 1.14 | 1.08 | 1.20 | 1.13 | 1.07 | 1.20 |
| Proportion students | 0.88 | 0.84 | 0.92 | 0.92 | 0.82 | 1.01 | 0.88 | 0.83 | 0.93 | 0.89 | 0.83 | 0.94 |
| Care home in area | 0.91 | 0.82 | 1.01 | 0.78 | 0.62 | 0.99 | 0.95 | 0.84 | 1.08 | 1.01 | 0.89 | 1.14 |

*Model fit (log marginal-likelihood)* – Model for period 6$^{th}$ Nov – 13$^{th}$ Jan = -969; Model for period 6$^{th}$ Nov – 2$^{nd}$ Dec = -613; Model for period 3$^{rd}$ Dec – 5$^{th}$ Jan = -732; Model for period 6$^{th}$ Jan – 31$^{st}$ Jan = -800.

**Appendix D: Sensitivity analysis – alternative measures of internet usage**

Our area level of digital inequality, internet user classification (Table 1), is useful since it is a multi-dimensional measure that combined information across a range of measures about confidence in using internet technologies. Key variables included in the construction of the classification of areas includes whether individuals own internet-enabled technologies, internet speed, how frequent individuals use the internet by different modes (e.g. laptop or mobile phone), information seeking behaviours online, use of social media or communication technologies, and online shopping habits. Descriptions of the area types are presented in Table D1.

**Table D1: Description of Internet User Classification area types.**

| Area type | Description |
|---|---|
| e-Cultural Creators | High levels of internet use, especially for social media, communication, streaming and gaming. Typically younger and student populations. |
| e-Professionals | High levels of engagement and experienced users, especially for online shopping. Urban professionals and high educational attainment. |
| e-Veterans | Frequent use of internet technologies across multiple devises and uses. Typically more affluent suburbs. |
| Youthful Urban Fringe | Primarily mobile users or use in public places. Higher levels of communication. Young and ethnically diverse populations. |
| e-Rational Utilitarians | High demand, but with poor infrastructure (often due to rural areas). Low levels of mobile use, with higher personal computers. Often middle aged or older adults. |
| e-Mainstream | Average characteristics for most measures and reflect diverse populations |
| Passive and Uncommitted Users | Limited or no interactions with the internet. Often from suburbs or rural areas. |
| Digital Seniors | Average use of internet, often from personal computers. Mostly older adults. |
| Settled Offline Communities | Limited engagement of the internet, with poor access to infastructure. Online shopping when use services. Retired people in rural areas. |
| e-Withdrawn | Least engaged with the internet, with lowest access and use. Deprived populations. |

Note: Descriptions taking from Singleton A, Alexiou A, Savani R. Mapping the geodemographics of digital inequality in Great Britain: An integration of machine learning into small area estimation. *Comput Environ Urban Syst* 2020;**82**:101486.

Internet User Classification is only a proxy measure of internet usage and its combination of variables may obscure specific associations. Here we re-run the analyses using a direct measure of data usage – median data usage (in gigabytes) for September 2019 produced by Ofcom (https://www.ofcom.org.uk/research-and-data/data/opendata). We suggest that these data are interpreted carefully, since median data usage in an area is not specifically confidence in using internet technologies (i.e. use is not necessarily the same as confidence, and skewed by streaming or downloads). Methods and data otherwise remain the same as presented in the main model. For brevity, we only present the results for the overall time period models (6th November 2020 to 31st January 2021). Continuous variables were centred and scaled to standardise them.

Table D2 presents summary statistics for each model. Standardised median data usage was negatively associated to both uptake (Relative Risk (RR) = 0.96, 95% Credible Intervals (CIs) = 0.92-0.99) and multiple tests (RR = 0.96, 95% CIs = 0.91- 1.00). The results suggest that in areas with greater internet usage, we observed fewer tests overall and fewer people who received multiple tests. Specifically, one standard deviation increase in the median data usage of an area would be associated with 4% fewer people having tests and 5% fewer people getting repeat tests. The finding is inconsistent with the result in the main analysis, and indeed our hypothesised directions.

**Table D2: Relative risks (RR) for the associations between covariates for each outcome (6th November 2020 to 31st January 2021). Note: Lower and upper limits refer to 95% Credible Intervals. Bold results have credible intervals that do not contain 1.**

| | **Outcome: overall uptake (persons)** | | | **Outcome: multiple tests (persons)** | | |
|---|---|---|---|---|---|---|
| | **RR** | **Lower** | **Upper** | **RR** | **Lower** | **Upper** |
| Deprivation score | 0.82 | 0.79 | 0.85 | 0.74 | 0.71 | 0.78 |
| Proportion of students in area | 0.95 | 0.92 | 0.98 | 0.92 | 0.88 | 0.96 |
| Care home beds / population | 1.16 | 1.08 | 1.25 | 1.19 | 1.08 | 1.31 |
| Average walking distance to nearest test site | 0.95 | 0.92 | 0.98 | 0.95 | 0.91 | 0.99 |
| Median data usage (GB) | 0.96 | 0.92 | 0.99 | 0.95 | 0.91 | 1.00 |

*Model fit (log marginal-likelihood)* – Model for uptake = -1971; Model for multiple uptake = -1765.

We also tested whether there was an interaction effects between digital inequality and neighbourhood deprivation to assess whether the association between degree of deprivation and the outcome change with digital inequality. This was because these two issues are inter-related, with deprived communities often less able to afford internet related technologies. For example, the correlation between neighbourhood level deprivation score and median data usage in Liverpool was 0.47. Inclusion of interaction effects for models with both median data usage and internet user classification fit separately did not improve model fit, with interaction effects non-significant. The high correlation between deprivation and median data usage may partly explain the lack of association in the expected direction for median data usage, with the association reflecting the residual effect of deprivation. However, we also note that IUC is also correlated to deprivation.

**Appendix E: Sensitivity analysis – individual level models**

One limitation of ecological analyses is the ecological fallacy where inferences about relationships at the area level cannot be made for individuals. This is important where we are interested in how *people* respond to asymptomatic testing, rather than how *communities* do. To extend the main analyses presented in the paper, we re-ran our models at the individual level. We present two models – one model for the likelihood of an individual having a positive test over the study period and one model for the likelihood of an individual having multiple tests over the study period. We were unable to undertake a similar analysis for lateral flow uptake as we did not have access to a full population register to identify who had received a test or not.

Multi-level binomial (binary) regression models were used to analyse each of our outcomes. The strength of the approach is the ability to independently test how individual level characteristics (level 1) and area level covariates (level 2) were associated to our outcome. Models were run using the same area level covariates as presented in the main paper, with age, sex and ethnic group included as individual-level covariates. Area level continuous variables were standardised (centred and scaled using z-scores). Sample size was 212 899 individuals nested within 298 LSOAs. Table E1 presents the model summary for both outcome measures. We divide the interpretation of the results by outcome measure. Relationships to likelihood of having had multiple (more than one) lateral flow test were often in the opposite direction compared to relationships to a positive test, suggesting that those populations less engaged with testing were also more likely to have benefitted from it.

Age was negatively associated to likelihood of having multiple lateral flow tests, suggesting that older adults were less likely to have multiple tests. Males were less likely to have had multiple tests than compared to females. Each ethnic group, other than 'Mixed', were also less likely to have received multiple tests than compared to the 'White' reference group. Deprivation was negatively associate to likelihood of having multiple tests, suggesting that individuals who lived in highly deprived areas were less likely to have had repeated testing. Individuals who lived in areas with student populations were less likely to have had multiple tests. There were no associations for care homes and access to test sites. Internet User Classification displayed several negative associations, suggesting that individuals who lived in areas characterised by communities less confident in using internet technologies were less likely to have received multiple tests.

For risk of having a positive test during the study period, age was negatively associated to the likelihood of a positive test. This would suggest that older adults were less likely to have tested positive. Males were more likely to have had a positive test compared to females. Few associations were detected by ethnic group, although individuals who were categorised in the 'Other' ethnic group were more likely to have had a positive test than compared to the 'White' reference group. In the area level covariates, deprivation was positively associated to likelihood of a positive test suggesting people who resided in deprived communities were more likely to have tested positive. Presence of a care home in an area was negatively associated to positivity, suggesting that individuals who lived in an area with a care home were less likely to have tested positive. Individuals who lived in areas with higher proportions of students were negatively associated to likelihood of a positive test.

**Table E1: Summary results from multi-level binomial regression exploring the factors associated with individual level likelihood of a positive test or whether an individual had multiple tests (both for any point between 6th November 2020 to 31st January 2021). Note: Std. Error = Standard Error. Estimates for regression coefficients are log odds. Estimates for random effects represent the variance and standard deviation.**

| | **Outcome: Multiple LFTs** | | | **Outcome: Positive test** | | |
|---|---|---|---|---|---|---|
| | **Estimate** | **Std. Error** | **p-value** | **Estimate** | **Std. Error** | **p-value** |
| Individual level covariates | | | | | | |
| Age | -0.003 | 0.0002 | <0.001 | -0.007 | 0.001 | <0.001 |
| Sex: | | | | | | |
| Female | Reference | | | Reference | | |
| Male | -0.180 | 0.009 | <0.001 | 0.120 | 0.030 | <0.001 |
| Ethnic group: | | | | | | |
| White | Reference | | | Reference | | |
| Asian | -0.363 | 0.026 | <0.001 | -0.056 | 0.085 | 0.512 |
| Black | -0.216 | 0.031 | <0.001 | 0.155 | 0.090 | 0.085 |
| Mixed | -0.036 | 0.037 | 0.325 | -0.144 | 0.130 | 0.269 |
| Other | -0.416 | 0.047 | <0.001 | 0.616 | 0.107 | <0.001 |
| Area level covariates | | | | | | |
| Deprivation | -0.114 | 0.020 | <0.001 | 0.113 | 0.023 | <0.001 |
| Proportion of students in area | -0.074 | 0.019 | <0.001 | -0.168 | 0.029 | <0.001 |
| Care home | 0.033 | 0.024 | 0.177 | -0.134 | 0.055 | 0.015 |
| Average distance to nearest test site | -0.004 | 0.010 | 0.665 | | | |
| Internet User Classification: | | | | | | |
| e-Veterans | Reference | | | | | |
| Digital Seniors | -0.115 | 0.079 | 0.144 | | | |
| e-Cultural Creators | -0.178 | 0.080 | 0.026 | | | |
| e-Mainstream | -0.106 | 0.045 | 0.019 | | | |
| e-Professionals | -0.146 | 0.056 | 0.010 | | | |
| e-Rational Utilitarians | -0.051 | 0.078 | 0.512 | | | |
| e-Withdrawn | -0.238 | 0.063 | <0.001 | | | |

| | | | | | |
|---|---|---|---|---|---|
| Passive and Uncommitted Users | -0.185 | 0.051 | <0.001 | | |
| Settled Offline Communities | -0.081 | 0.126 | 0.517 | | |
| Youthful Urban Fringe | -0.154 | 0.070 | 0.028 | | |
| <u>Random effects (variance and standard deviation)</u> | | | | | |
| Lower Super Output Area | 0.024 | 0.154 | | 0.091 | 0.302 |
| <u>Model fit</u> | | | | | |
| AIC | 287026 | | | 44650 | |
| BIC | 287242 | | | 44763 | |
| Deviance | 286984 | | | 44628 | |
